# LITERATURE REVIEW: DISASTER RISK REDUCTION PROGRAMS TO INCREASE PUBLIC AWARENESS OF NATURAL DISASTERS

**DOI:** 10.1101/2023.12.15.23300051

**Authors:** Susi Wahyuning Asih, Moses Glorino Rumambo Pandin, Ahmad Yusuf, Supriyadi

## Abstract

**Background:** The key components of disaster risk reduction typically include risk assessment, early warning systems, public awareness and education, infrastructure and land-use planning, preparedness and response planning, and sustainable development. The third component is the public awareness and education. Educating communities about potential risks and how to prepare for and respond to disasters is crucial for building resilience and ensuring the effective implementation of DRR measures. The fourth component is infrastructure and land-use planning. Regarding the preparedness and response planning, developing comprehensive disaster preparedness and response plans is helpful in ensuring a swift and coordinated response during emergencies, thereby minimizing the impact on life and property. Lastly, regarding sustainable development, integrating DRR into development planning can help create sustainable and resilient communities that are better equipped to withstand and recover from disasters.

**Purpose:** This study aims to review articles that examine the disaster risk reduction program to increase public awareness of natural disasters

**Design:** This study was categorized as a literature review

**Method:** Data were collected by searching articles published on SAGE, Springer link, Proquest, Scopus, and Science Direc in 2020-2023.

**Results:** 263 articles were used in the review. These articles discuss the disaster risk reduction program to increase public awareness of natural disasters. Fifteen articles reviewed were original research.

**Conclusion:** Local communities play a central role in hazard identification, development of preparedness plans, detection and response to emergencies, and implementation of recovery efforts. Community leaders and local health workers (e.g. family doctors, nurses, midwives, pharmacists, community health workers).

## BACKGROUND

Disaster management seen from the perspective of community perception is very urgent and important to know as basic knowledge for the community about what to do in the face of disasters. The main components of disaster risk reduction (DRR) usually include disaster risk assessments, early warning systems, community awareness and education, and sustainable development. Risk assessment involves identifying, assessing, and analyzing the risks associated with various hazards, including their potential impact on society and the environment. Meanwhile, early warning systems may help minimize the impact of disasters by alerting populations of imminent catastrophes in a timely and precise manner. (M.A. Chisty,2020). Regarding the third component, i.e., community awareness and education, educating communities about potential risks and how to prepare for and respond to disasters is critical to building resilience and ensuring effective implementation of DRR measures. Lastly, integrating DRR into development planning can help create sustainable and resilient communities that are better prepared to survive and recover from disasters.

Disasters are no longer considered a sporadic phenomenon but are managed and reduced as much as possible. It is no longer considered a danger that is impossible to handle. However, The issue of how to include the concept of safety in a catastrophic event requires further studies. To include institutions and social units in the field of catastrophe studies, the concept of “vulnerability” emerges, particularly concerning the existence of civilization and human life on the planet. Disasters, their causes, and their effect on human existence must be minimized and identified as early as possible to facilitate the development of a solution for disaster management (H. Setyawan, 2021).

Several studies have shown that treatments based on lifestyle might prevent between 40 and 70 percent of the implementation of a Disaster Risk Reduction (DRR) program. Disaster risk reduction in rural areas is necessary to increase the community’s ability to cope with the disaster-related risks. In Kanyasan’s study (2018), Community-based DRR, also known as the Community-Managed Disaster Risk Reduction training program, aims to empower the village community (Kanyasan K, 2018). Since the one-month training involves impacted village residents, it is extremely important to ensure the program significance. The initial hazard posed by the tragedy was landslides in the mountainous sections of the village, jeopardizing homes and agricultural land. This was the first step in gathering community members impacted by the disaster. Floating water is the second potential catastrophic concern.

Landslides occurring in the steep areas of the village pose the greatest risk to the settlements and agricultural land. This is the first hazard that might bring about a potential calamity. Floods are the second type of natural catastrophe that might occur. Floods that strike low-lying village sites, agricultural land, and settlements require good mapping (Haque A, 2022). The absence of a village spatial planning policy or long-term development plan for managing natural resources causes frequent shifts to land use, eventually resulting in shrinkage of agricultural land and increased risk of floods. Overstress on regional ecosystems and a decline in environmental carrying capacity are consequences of the population’s increasingly pervasive poverty. Disaster risk management (DRM) expertise is still lacking among village residents and their elected officials. Additionally, women are underrepresented in the decision-making and strategic policy-making processes related to disaster management and response. Many of the village’s religious and social groups are now disorganized, but they potentially become a powerful voice in the DRR disaster risk management campaign (Lawangen A, 2023).

The village exhibits a strong culture of cooperation, tolerance, and the spirit of self-reliance, which are important factors in disaster management. Several steps are required to increase the disaster risk management capacity of village communities affected by disasters, **the first step** is to develop the community’s understanding of the threats, vulnerabilities, capacities, and potential risks of landslides, floods, and factors that influence their area. **Secondly**, it is necessary to prepare potential local cadres to mobilize community participation in initiatives aimed at reducing disaster risks. **The third step is to** carry out participatory risk mapping of landslides and floods. **The fourth step** is to amplify women’s role and strategic positions in village disaster risk management activities. Lastly, it is necessary to develop innovative actions for disaster risk management in village communities.

### Method

A literature study was conducted by scanning three databases (SAGE, Springer Link, Proquest, Scopus, and Science Direct) for relevant peer-reviewed articles published in English between 2020 and 2023. This review specifically targeted studies focusing on how DRR is put into practice. Encompassing experimental design, observation, and randomized and non-randomized controlled trials, the selected studies focused on the community’s efforts to reduce catastrophe risk. The DRR’s output was the primary outcome. The literature search only included open-access papers published between 2020 and 2023.

### Result

The author found 31,232 articles from three databases, including Scopus: 689 articles; Pubmed: 15,766 articles; CINAHL: 14,777 articles, after searching with specific keywords combined with Boolean “AND” and “OR” operators. The keywords of the PICO framework include Population (communities), Intervention (Mitigation Disaster), Comparison (cognitive and psychomotor), and Results (Disaster Risk Reduction condition).

Researchers also used this literature review search criteria with the PRISMA flow diagram to obtain appropriate and eligible research articles. The following is a diagram image *flow* PRISMA:

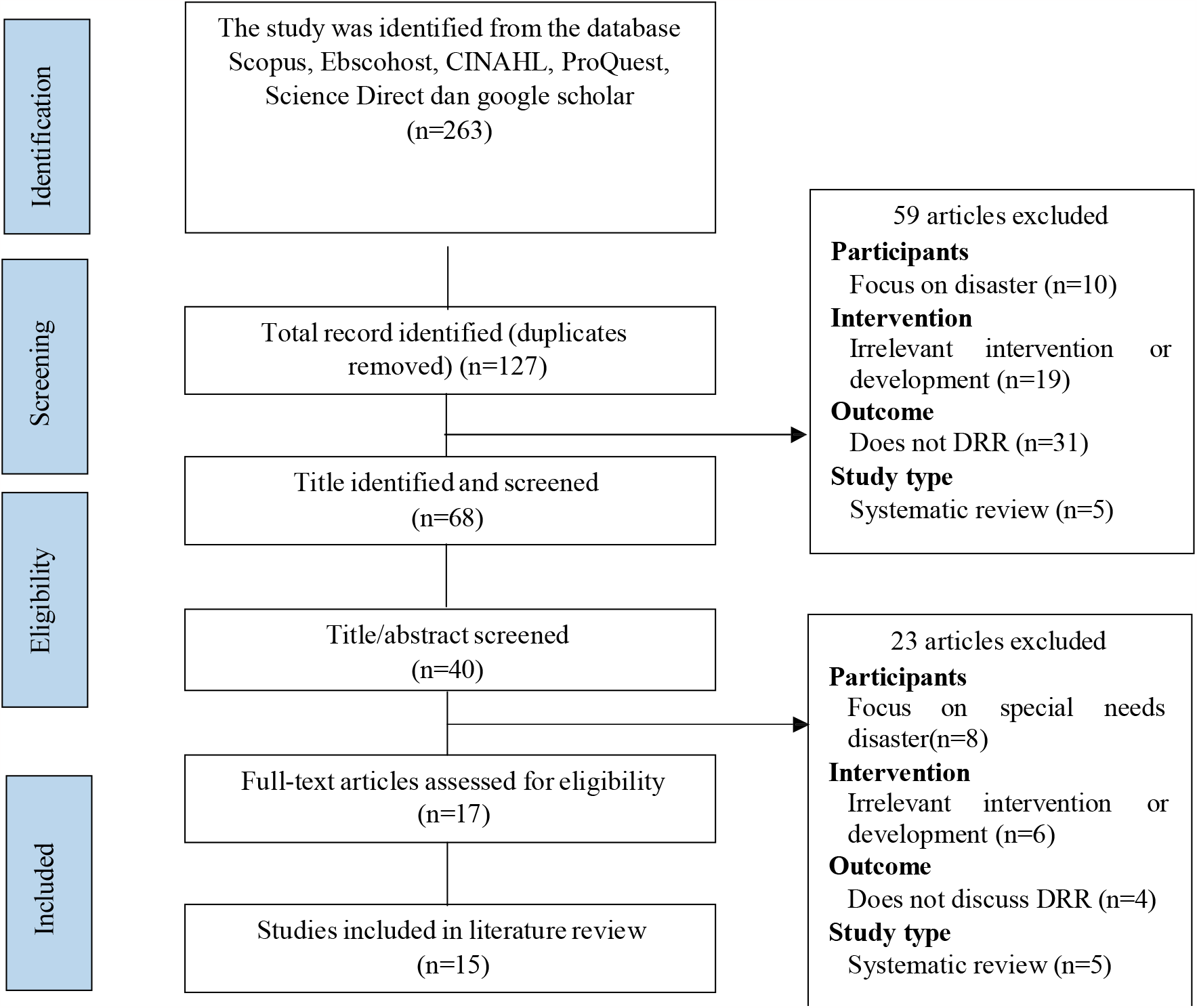

**Tabel 1.**
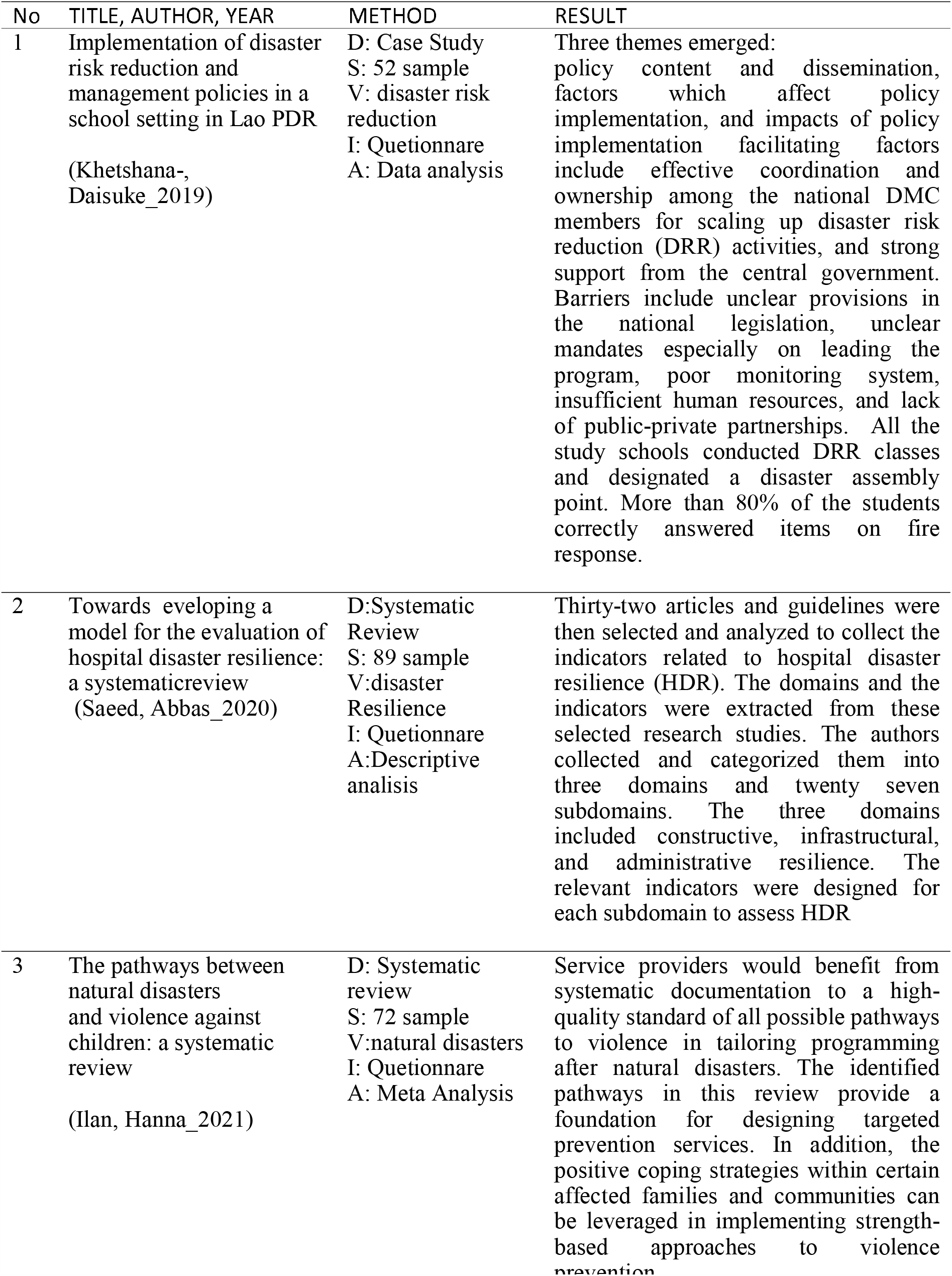
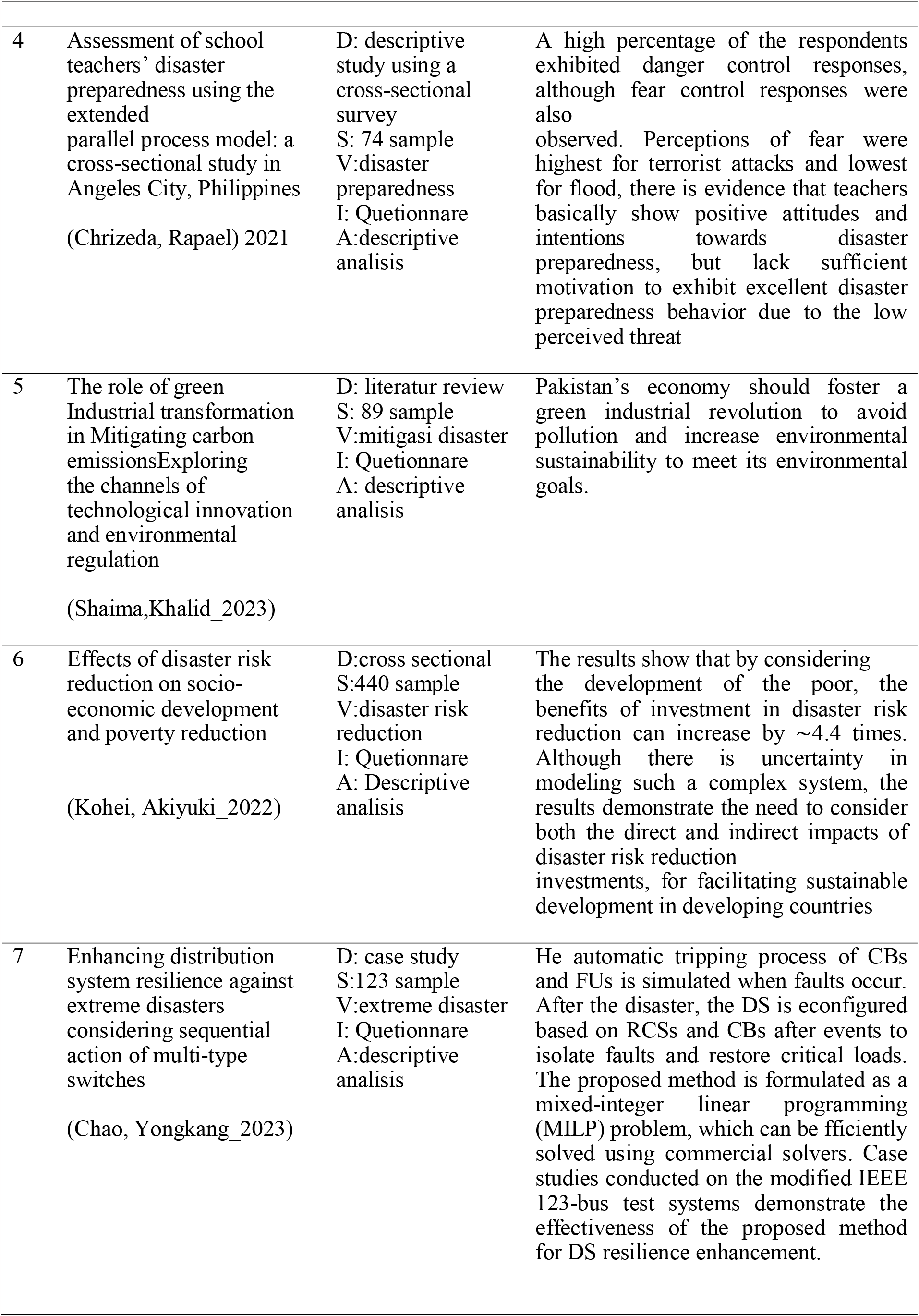
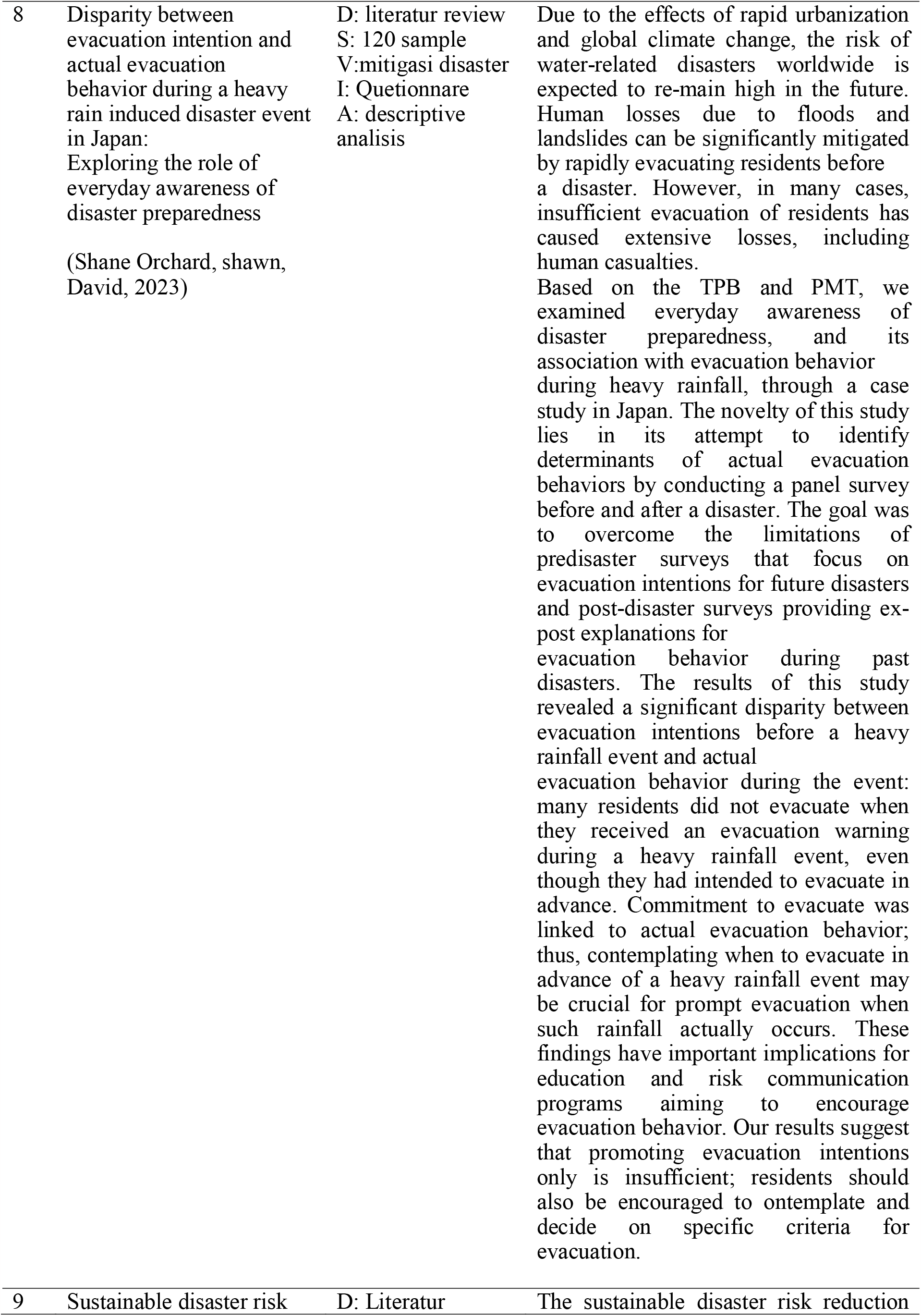
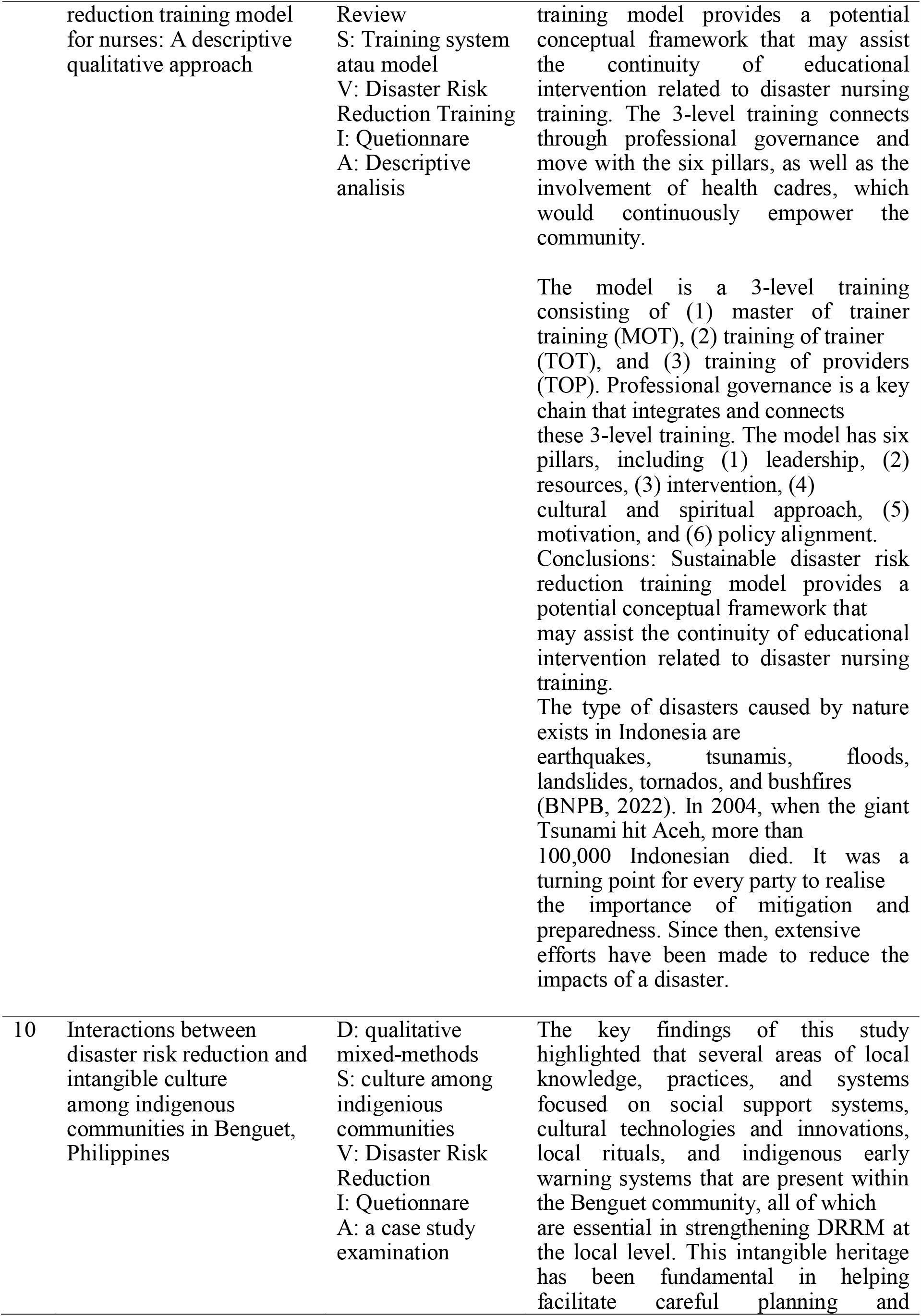
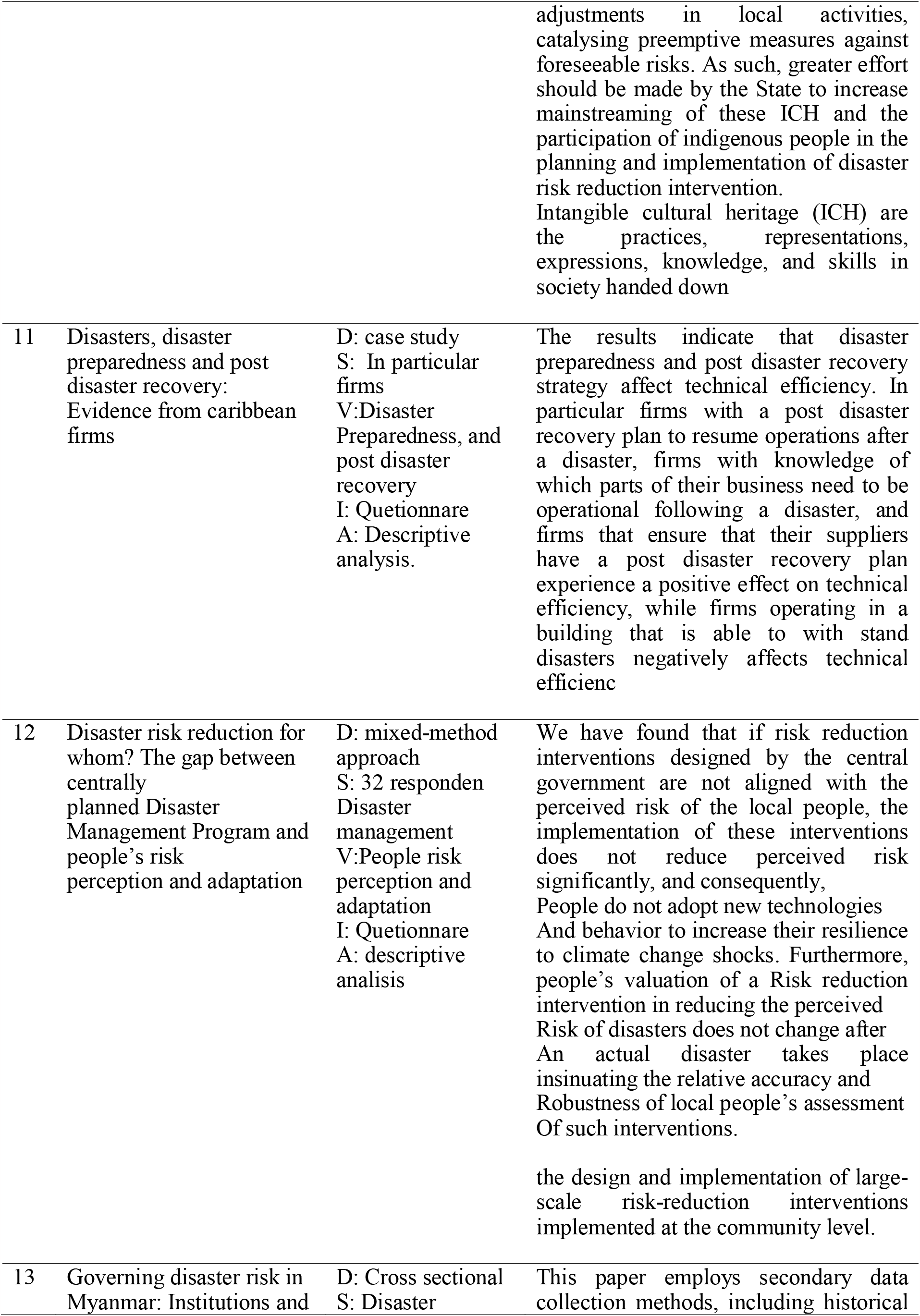
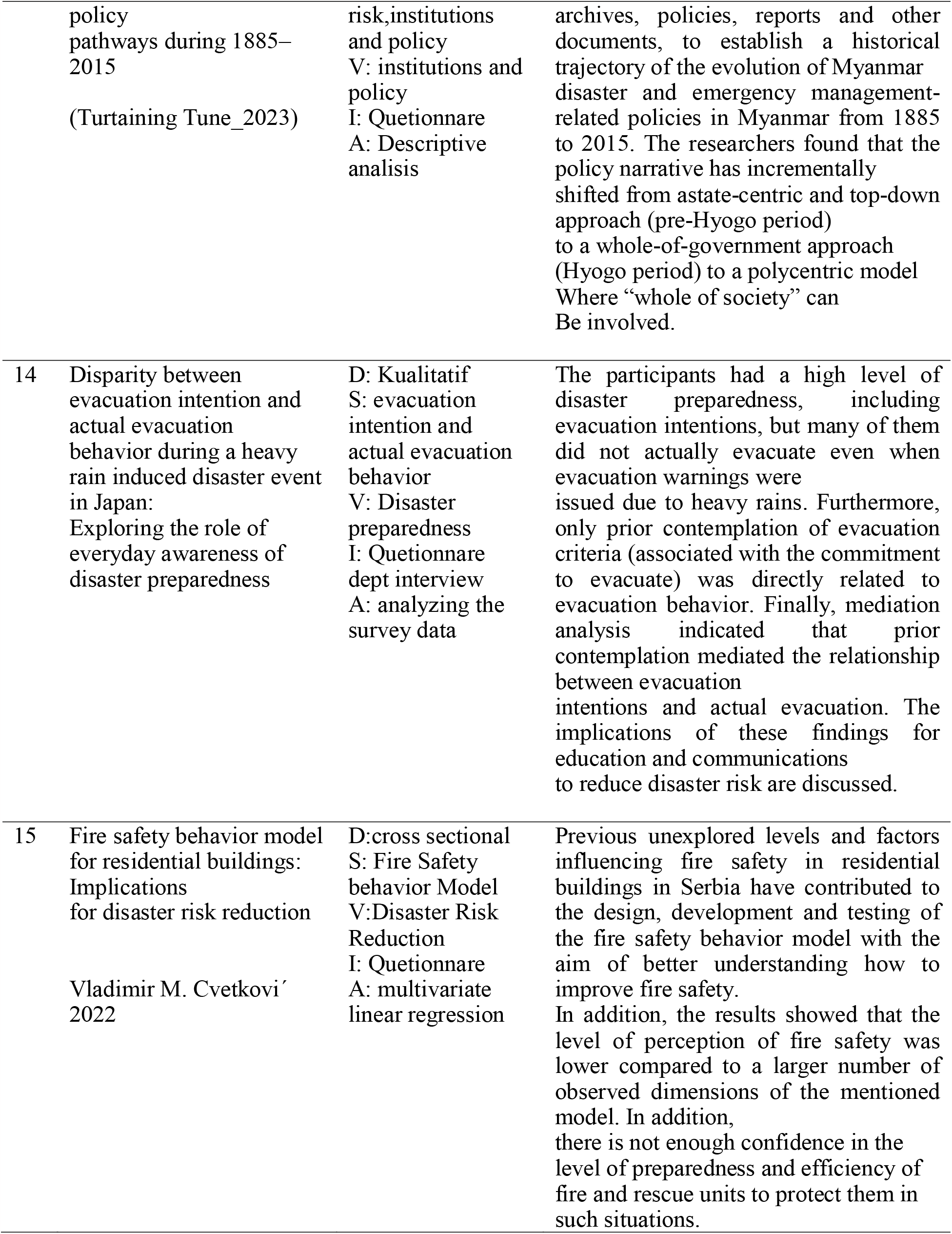
Characteristics Of Review Studies.

## Discussion

### Disaster Risk Reduction

A disaster is defined as an event or sequence of events that endangers and interrupts human life and economic activity due to human action or inaction, and which causes physical harm to the environment, loss of property, and emotional distress (Law No. 24/2007 Concerning Disaster Management). The International Strategy for Disaster Reduction (ISDR) defines disaster as events that significantly impair a society’s functioning, large-scale human casualties due to material, economic, or environmental factors, and surpass the capacity of the affected community to recover using their own resources (Okuda K, Kawasaki, 2022).

### Disaster Mitigation

When discussing about mitigation, it involves reducing risks to minimize loss. The mitigation process encompasses a variety of activities and measures. Possible first steps toward safety include making plans ahead of time, assessing the danger level, and implementing a disaster management plan that incorporates rescue, rehabilitation, and relocation (Mansourian, 2021). According to Decree no. 131 of 2003, which is signed by the minister of home affairs of the Republic of Indonesia, the term “mitigation” is defined as the measures taken to lessen the impact of natural catastrophes. Important aspects of mitigation include being well-prepared, watchful, and equipped with a range of skills to deal with disasters.

Various initiatives to increase public knowledge and preparedness in anticipating disasters have been initiated in Indonesia at various administrative levels. This effort involves many agencies and institutions at the local national, and international levels (Preeya S.Mohan, 2023). However, the effectiveness of these efforts is still unclear because there is no framework and tools to assess the level of community preparedness and the sustainability of these activities. So far, efforts to increase community preparedness have been carried out based on the concerns, objectives, and capabilities of the institutions concerned. Therefore, it is still uncertain whether these efforts have fulfilled all the elements required for disaster preparedness.

### Safety of Disaster

The paradigm has shifted from perceiving disasters as random occurrences to actively engaging in mitigation and controlling them whenever possible. The most important issue, however, is how to include safety measures in this catastrophe. For social organizations and units to fit in with society, the concept of “vulnerability” evolved. Natural disasters can occur at any time, is often unpredictable, and catch people off guard, leading to a lack of preparedness when a disaster strikes (Lukewich J, 2023). This unpreparedness may be accounted for by insufficient knowledge regarding preparedness in facing disasters. It is necessary to prepare the community through activities that prepare them to anticipate disaster quickly and effectively to protect them from significant losses in disasters. Disaster readiness is the state in which society, both at an individual and collective level, has the physical and psychological capacity to deal with the aftermath of a catastrophe.

According to Benjamin Blum, the knowledge or cognitive aspect plays a pivotal role in determining an individual’s actions. Blum’s theory posits that, in addition to environmental factors, people’s attitudes and actions have the greatest influence on individuals and communities. Environmental variables are ranked as the top factor according to this viewpoint. There is a strong correlation between one’s attitude and their perception, personality, and motivation, making attitude a key factor in behavior modification. A person’s attitude might be defined as the conceptual framework they develop through their life experiences. A person’s reactions to other people, objects, and situations in his life are influenced by it (Hatori T, 2023). Attitudes inherently include emotional components, cognitive components (including views, ideas, and beliefs), and behavior, and perceptions are inextricably linked to this.

A person’s attitude, as defined by Sukidjo, is a state of mind and nervous system preparedness influenced by experience, directly affecting how individuals respond to everything that happens to them. An individual’s moral stance represents their evaluation of any given situation or item. After becoming familiar with the stimuli or item, individuals will likely evaluate or act in response to it. Management of disaster emergency response includes post-disaster reconstruction or rehabilitation by paying attention to local wisdom and increasing community capacity.

Urgent disaster risks can be addressed by conducting disaster management training or simulations to improve the public’s understanding of dealing with disaster risks. Disasters are no longer considered a sporadic phenomenon but as manageable and reducible events. However, how do we then place the safety element in this disaster? The emergence of the idea of “vulnerability” is developed to accommodate institutions and social units as parts.

## CONCLUSION

Local community plays an important role in identifying hazards, designing preparation plans, detecting and responding to crises, and executing recovery operations. Leaders in the community and local health care providers (such as family physicians, nurses, midwives, pharmacists, and community health workers) have the power to establish credibility, spread knowledge, and identify vulnerable individuals.

Disaster risk reduction is the concept and practice of systematically reducing disaster risk. Some examples of the efforts include reducing the factors that cause exposure to hazards, reducing human and property vulnerability, managing land and the environment, and increasing preparedness for disaster impacts, among others. Disaster risk reduction includes disaster management disaster mitigation and disaster preparedness, and is also a part of sustainable development. Disaster risk reduction efforts are very important in every aspect of life. Although disaster events are unavoidable, it is very important to optimize risk reduction efforts so that when a disaster occurs, society can immediately bounce back and recover quickly.

## Data Availability

All data produced in the present study are available upon reasonable request to the authors

## ACKNOWLEDGMENTS

Thank you to the nursing lecturers at Airlangga University and friends who have helped write this article.

